# Impact of the COVID-19 nonpharmaceutical interventions on influenza and other respiratory viral infections in New Zealand

**DOI:** 10.1101/2020.11.11.20228692

**Authors:** Q. Sue Huang, Tim Wood, Lauren Jelley, Tineke Jennings, Sarah Jefferies, Karen Daniells, Annette Nesdale, Tony Dowell, Nikki Turner, Priscilla Campbell-Stokes, Michelle Balm, Hazel C Dobinson, Cameron C. Grant, Shelley James, Nayyereh Aminisani, Jacqui Ralston, Wendy Gunn, Judy Bocacao, Jessica Danielewicz, Tessa Moncrieff, Andrea McNeill, Liza Lopez, Ben Waite, Tomasz Kiedrzynski, Hannah Schrader, Rebekah Gray, Kayla Cook, Danielle Currin, Chaune Engelbrecht, Whitney Tapurau, Leigh Emmerton, Maxine Martin, Michael G. Baker, Susan Taylor, Adrian Trenholme, Conroy Wong, Shirley Lawrence, Colin McArthur, Alicia Stanley, Sally Roberts, Fahimeh Ranama, Jenny Bennett, Chris Mansell, Meik Dilcher, Anja Werno, Jennifer Grant, Antje van der Linden, Ben Youngblood, Paul G. Thomas, Richard J. Webby, on behalf of the investigation team

## Abstract

Stringent nonpharmaceutical interventions (NPIs) such as lockdowns and border closures are not currently recommended for pandemic influenza control. New Zealand used these NPIs to eliminate coronavirus disease 2019 during its first wave. Using multiple surveillance systems, we observed a parallel and unprecedented reduction of influenza and other respiratory viral infections in 2020. This finding supports the use of these NPIs for controlling pandemic influenza and other severe respiratory viral threats.

## Introduction

The coronavirus disease 2019 (COVID-19) pandemic, declared by the World Health Organisation (WHO) on 11 March 2020, reached New Zealand (NZ) on 28 February 2020. From 2 February 2020, NZ introduced progressive border restrictions and a 4-level alert system aiming to eliminate COVID-19.^1^ Soon after the emergence of community transmission of COVID-19, the stringent nonpharmaceutical interventions (NPIs) of Level-4 (nationwide lockdown) were applied, lasting from 25-March to 27-April-2020. These included: 1) blocking importation of the virus (border closure to non-New Zealanders, and 14-day quarantine for returning travellers); 2) stamping out transmission within NZ (widespread testing, isolating cases, contact tracing and quarantine of exposed persons); 3) physical distancing measures (stay-at-home orders, cancelling all gatherings, closing schools, non-essential businesses and all public venues, and restricting domestic travel); 4) individual infection prevention and control measures (promoting hand hygiene and cough etiquette, staying home with mild respiratory symptoms, and mask wearing if unwell); and 5) communicating risk to the public and various stakeholders. The implementation of these NPIs combined with public compliance effectively eliminated community transmission of COVID-19 during the first wave (12-February to 13-May), achieving 101 consecutive days without detection of community COVID-19 cases.^2,3^ Since this implementation, NZ has continued to apply NPIs in various forms up until submission of this report.^1^

The effectiveness of NPIs in reducing viral transmission depends on transmission characteristics of the virus.^4^ If a substantial proportion of transmission occurs before the onset of symptoms (i.e. pre-symptomatic shedding) or during asymptomatic infection, the population impact of health screening and case-patient isolation will be diminished.^5^ Influenza virus has a shorter serial interval and earlier peak infectivity compared to SARS-CoV-2. Our recent publication also showed that up to 32% of influenza virus infections in NZ are mild or asymptomatic, suggesting the likelihood of substantial asymptomatic transmission.^6^ These characteristics led to the assumption that these NPIs would be less successful in controlling influenza virus than coronaviruses,^7^ however, robust field data are lacking. New Zealand’s use of stringent NPIs created a natural experiment enabling an understanding of the impact of these NPIs on illnesses caused by influenza and other respiratory viruses. This type of knowledge is valuable for informing pandemic influenza preparedness and seasonal influenza planning for the northern hemisphere’s upcoming winter in the context of the ongoing COVID-19 pandemic.

Here we describe the complete absence of the usual winter influenza virus epidemic and a remarkable reduction of other respiratory viral infections in NZ during and after the implementation of these stringent NPIs in 2020.

## Results

Influenza activity in NZ during the winter of 2020 was very low as confirmed by multiple national surveillance systems (Figure 1).

**Figure 1.**
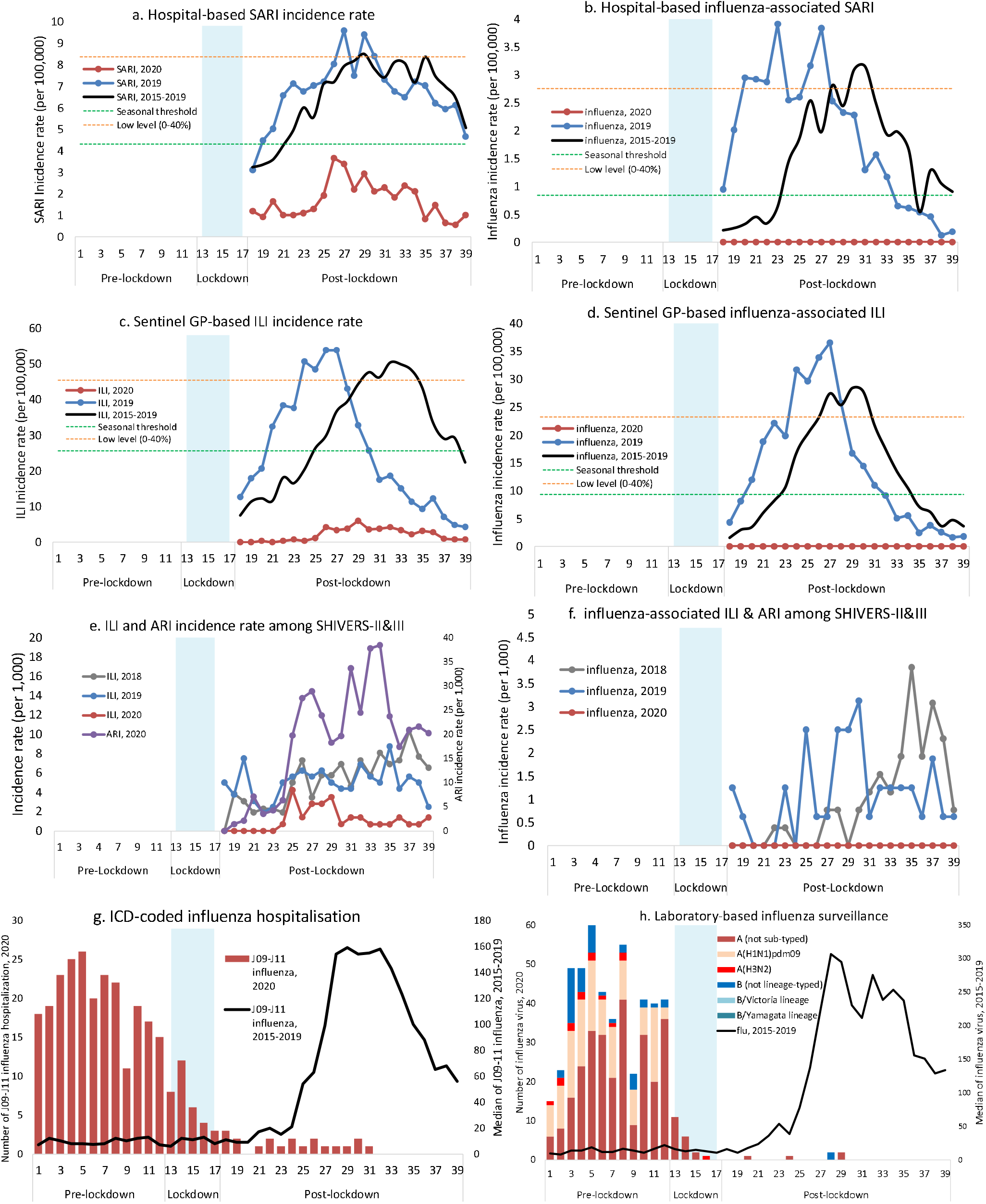
Temporal distribution of acute respiratory illness and associated influenza infections in 2020 compared with previous years Abbreviations: SARI (severe acute respiratory illness); GP (general practice); ILI (influenza-like illness); ARI (acute respiratory illness); SHIVERS-II&III (The second and third iterations of the Southern Hemisphere Influenza and Vaccine Effectiveness Research and Surveillance programme); ICD (International Classification of Diseases); flu refers to influenza. The calculation for epidemic threshold and low influenza activitiy is described in the Methods. A patient with cough and history of fever (subjective fever or measured temperature ≥ 38°C) and onset within the past 10 days meets the SARI case definition if hospitalised or meets the ILI case defintion if consulting a GP or participating in the SHIVERS-II&III study. The ARI case definition among SHIVERS-II&III participants refers to an “acute respiratory illness with fever or feverishness and/or one of following symptoms (cough, running nose, wheezing, sore throat, shortness of breath, loss of sense of smell/taste) with onset in the past 10 days.”

From May to September 2020, hospital-based severe acute respiratory illness (SARI) surveillance (catchment population of 1 million people) showed very low SARI incidence rates, all below the seasonal threshold defined by the reference period of 2015-2019 (F1-a). No influenza-associated SARI were identified (F1-b).

The national sentinel general practice (GP)-based surveillance usually covers ∼10% of the NZ population and captures patients with influenza-like illness (ILI) attending medical consultations. However, this patient flow was altered in 2020 as many patients with ILI were channelled to COVID-specific testing centres where patients were predominantly only tested for SARS-CoV-2. Additionally, the number of participating practices were 18-57% lower than the usual participation rate over the surveillance period. The ILI incidence rates were below the seasonal threshold compared to the reference period (F1-c). No influenza-associated ILI were detected (F1-d). Independently, the low ILI incidence rates were also observed in the HealthStat GP-based ILI surveillance system (Supplementary Figure 1).

SHIVERS-II & III (the second and third iterations of the Southern Hemisphere Influenza and Vaccine Effectiveness Research and Surveillance programme) are two community-based cohorts that follow ∼1400 adults aged 20-69 years and ∼80 infants in the Wellington region, respectively. Active surveillance for both cohorts in 2020 included swabbing of participants meeting the case definition for ILI and/or acute respiratory illness (ARI). The ILI incidence rate in 2020 was lower than the previous years of 2019 and 2018, however, ARI incidence was high (F1-e). No influenza-associated ILI or ARI were identified (F1-f).

The national International Classification of Diseases (ICD)-coded (ICD-10AM-VI code J9-J11) influenza hospitalisations for all NZ public hospitals showed a significant decline (F1-g). From 1-January to 31-July-2020, a total of 291 influenza hospitalisations were coded: pre-lockdown 238 (81.8%); lockdown 33 (11.3%); and post-lockdown 15 (5.2%). The Cochran–Armitage test showed a significant downward trend (p<0.001) in influenza hospitalisations.

The laboratory-based surveillance system includes testing samples ordered by clinicians during routine clinical diagnostic processes for hospital inpatients and outpatients (serving ∼70% of the NZ population). Additionally, this system also includes testing samples from all influenza surveillance systems (F1-h). During the COVID-19 laboratory response, some labs may have prioritised testing for SARS-CoV-2 over influenza and other respiratory viruses. From 1-January to 27-September-2020, there were 500 influenza virus detections: pre-lockdown 474 (94.8%); lockdown 20 (4.0%); and post-lockdown 6 (1.2%). The Cochran–Armitage test showed a significant downward temporal trend (p<0.001) in influenza virus detections.

Table 1 shows the number of respiratory viruses detected and the proportional reduction for each virus in 2020 (versus the reference period of 2015-2019) before, during and after the lockdown. Dramatic reductions were observed for influenza virus compared with the reference period: 67.7% reduction during and 99.9% after the lockdown. Marked reductions were also evident for other respiratory viruses during post-lockdown compared with the reference period (for temporal distribution, see supplementary Figure 2): respiratory syncytial virus (98.0% reduction), human metapneumovirus (92.2%), enterovirus (82.2%), adenovirus (81.4%), parainfluenza virus types 1-3 (80.1%), and rhinovirus (74.6%). During post-lockdown when the restrictions were eased to Level-1, we observed a significant increase in the proportion of rhinovirus compared to the median rate for this period from the preceding period: 33% (175/529) from 8-June to 11-August vs 4.8% (10/209) from 13-May to 7-June (p<0.0001). The rhinovirus-associated incidence rates in 2020 among SHIVERS-II&III and SARI surveillance also increased after the ease of restrictions (Supplementary Figure 3).

**Table.** The number of influenza and other respiratory viruses detected and their reduction in 2020 before, during and after the lockdown compared with the reference period of 2015-2019

| Virus | Pre-Lockdown |  |  |  | Lockdown | Post-Lockdown |  |  |  |  |  |
| --- | --- | --- | --- | --- | --- | --- | --- | --- | --- | --- | --- |
|  |  |  |  |  | Level 4 | Level 3 | Level 2 | Level 1 | Level 2&3 | Level 2&1 |  |
| Month | Jan | Feb | 1-24Mar | Subtotal | 25Mar-27Apr | 28Apr-12Ma | 13May-7Jun | 8Jun-11 Aug | 12Aug-29Aug | 31Aug-27Sept | Subtotal |
| No. weeks | 1 to 5 | 6 to 9 | 10 to 12 |  | 13 to 17 | 18 to 19 | 20 to 23 | 24 to 32 | 33 to 35 | 36 to 39 |  |
| Influenza |  |  |  |  |  |  |  |  |  |  |  |
| No. (2020) | 196 | 156 | 122 | 474 | 20 | 0 | 1 | 5 | 0 | 0 | 6 |
| Median (2015-19) | 84 | 55 | 62 | 206 | 62 | 27 | 126 | 2618 | 731 | 544 | 5062 |
| IQR (2015-19) | 39-85 | 30-94 | 21-66 | 90-240 | 51-139 | 20-79 | 89-318 | 1120-3165 | 549-996 | 513-935 | 3131-5097 |
| % Reduction* | -133.3 | -183.6 | -96.8 | -130.1 | 67.7 | 100.0 | 99.2 | 99.8 | 100.0 | 100.0 | 99.9 |
| RSV |  |  |  |  |  |  |  |  |  |  |  |
| No. (2020) | 9 | 16 | 9 | 34 | 16 | 3 | 8 | 15 | 5 | 3 | 34 |
| Median (2015-19) | 14 | 15 | 15 | 44 | 85 | 46 | 162 | 1176 | 270 | 159 | 1743 |
| IQR (2015-19) | 14-15 | 11-16 | 15-20 | 40-45 | 43-93 | 40-57 | 159-167 | 1056-1219 | 253-361 | 146-287 | 1591-2103 |
| % Reduction* | 35.7 | -6.7 | 40.0 | 22.7 | 81.2 | 93.5 | 95.1 | 98.7 | 98.1 | 98.1 | 98.0 |
| Rhinovirus |  |  |  |  |  |  |  |  |  |  |  |
| No. (2020) | 103 | 177 | 238 | 518 | 26 | 2 | 10 | 175 | 95 | 44 | 326 |
| Median (2015-19) | 72 | 79 | 90 | 232 | 144 | 79 | 209 | 529 | 175 | 305 | 1282 |
| IQR (2015-19) | 67-85 | 76-87 | 77-111 | 220-303 | 135-193 | 62-94 | 206-227 | 498-538 | 172-216 | 244-336 | 1277-1384 |
| % Reduction* | -43.1 | -124.1 | -164.4 | -123.3 | 81.9 | 97.5 | 95.2 | 66.9 | 45.7 | 85.6 | 74.6 |
| Enterovirus |  |  |  |  |  |  |  |  |  |  |  |
| No. (2020) | 34 | 30 | 23 | 87 | 3 | 0 | 5 | 33 | 9 | 14 | 61 |
| Median (2015-19) | 40.5 | 34 | 34 | 107.5 | 69 | 26.5 | 44 | 110.5 | 53.5 | 84 | 343 |
| IQR (2015-19) | 34-48 | 29-45 | 25-54 | 102-133 | 58-81 | 20-35 | 42-51 | 88-140 | 41-56 | 67-107 | 262-383 |
| % Reduction* | 16.0 | 11.8 | 32.4 | 19.1 | 95.7 | 100.0 | 88.6 | 70.1 | 83.2 | 83.3 | 82.2 |
| Adenovirus |  |  |  |  |  |  |  |  |  |  |  |
| No. (2020) | 54 | 54 | 37 | 145 | 9 | 3 | 12 | 39 | 25 | 24 | 103 |
| Median (2015-19) | 83 | 59.5 | 66.5 | 207 | 75 | 46.5 | 79 | 227.5 | 84.5 | 129 | 554 |
| IQR (2015-19) | 45-151 | 53-77 | 54-76 | 156-299 | 73-100 | 41-55 | 70-105 | 207-290 | 75-131 | 89-237 | 519-780 |
| % Reduction* | 34.9 | 9.2 | 44.4 | 30.0 | 88.0 | 93.5 | 84.8 | 82.9 | 70.4 | 81.4 | 81.4 |
| hMPV |  |  |  |  |  |  |  |  |  |  |  |
| No. (2020) | 11 | 26 | 45 | 82 | 10 | 3 | 10 | 17 | 7 | 3 | 40 |
| Median (2015-19) | 7 | 4 | 6 | 16 | 10 | 5 | 24 | 187 | 103 | 159 | 513 |
| IQR (2015-19) | 7-10 | 2-5 | 4-10 | 12-21 | 5-13 | 5-8 | 20-26 | 161-207 | 95-109 | 147-178 | 458-538 |
| % Reduction* | -57.1 | -550.0 | -650.0 | -412.5 | 0.0 | 40.0 | 58.3 | 90.9 | 93.2 | 98.1 | 92.2 |
| PIV |  |  |  |  |  |  |  |  |  |  |  |
| No. (2020) | 25 | 25 | 22 | 72 | 32 | 9 | 24 | 41 | 18 | 9 | 101 |
| Median (2015-19) | 27 | 9 | 7 | 43 | 28 | 17 | 31 | 230 | 82 | 148 | 508 |
| IQR (2015-19) | 24-28 | 8-12 | 6-8 | 38-45 | 21-37 | 16-19 | 31-55 | 200-317 | 66-148 | 136-231 | 398-803 |
| % Reduction* | 7.4 | -177.8 | -214.3 | -67.4 | -14.3 | 47.1 | 22.6 | 82.2 | 78.0 | 93.9 | 80.1 |

\*%Reduction = 1 - [No. virus (2020)/median no. virus (2015-2019)]%; IQR refers to interquartile range; RSV refers to respiratory syncytial virus; hMPV refers to human metapneumovirus; PIV refers to parainfluenza virus types 1-3
New Zealand COVID-19 alert levels and dates and related public health measures can be accessed at:
https://covid19.govt.nz/assets/resources/tables/COVID-19-alert-levels-summary.pdf

## Discussion

New Zealand, a southern hemisphere country with a temperate climate, has a well-established influenza circulation pattern with peak incidences in the winter months.^8-10^ Multiple surveillance systems showed that there was no annual laboratory-confirmed influenza outbreak or epidemic detected during the 2020 winter season. Remarkably, influenza virus circulation was almost non-existent during the 2020 winter, a 99.9% reduction compared with previous years. We postulate that NZ’s use of stringent NPIs (lockdowns and border controls) have markedly changed human behaviour, resulting in substantial reductions in contacts between influenza-infected individuals and influenza-susceptible individuals. The nationwide lockdown occurred during late autumn before the usual influenza season. This timing may also be important as the resulting small number of influenza-infected individuals did not appear sufficient to trigger a sustained influenza epidemic in the oncoming winter, in a setting of strict border control, personal hygiene promotion and ongoing forms of social distancing that remained in place after the lockdown.^1^

The WHO’s pandemic influenza intervention guidance does not recommend stringent NPIs when pandemic influenza reaches sustained transmission in the general population because these NPIs have been considered ineffective and impractical.^5^ However, the knowledge base used in developing WHO guidance for influenza pandemic prevention is limited and consists primarily of historical observations and modelling studies. In light of these data presented from NZ, previous experiences from Hong Kong during the 2003 SARS epidemic,^11^ and the COVID-19 pandemic,^12^ we suggest it is important to re-evaluate the role of NPIs in mitigating or even eliminating severe pandemic influenza. This knowledge may inform better preparedness for future influenza pandemics and seasonal influenza control and other severe respiratory viral threats.

Other potential contributing factors for the reduction in influenza virus detections include influenza vaccination, climatic changes and viral-viral interactions. The NZ National Immunisation Register recorded ∼22% influenza vaccine coverage in 2020 (35% more than recorded in the three months after the start of funded influenza vaccinations in 2020 compared with 2019, personal communication). Cold temperature promotes the ordering of lipids on the viral membrane which increases the stability of the influenza virus particle.^13^ Winter 2020 was NZ’s warmest winter on record. The nationwide average temperature was 9.6°C, 1.1°C above the 1981-2010 average.^14^ The warmer winter may reduce virus stability, contributing to lower influenza circulation. Rhinovirus, the most prevalent virus after the lockdown, may protect individuals against influenza virus infection. Interferon-stimulating immunity mediated by rhinovirus infection may make it difficult for additional viruses such as influenza to become established in a population.^15^ Similar observations were reported in Sweden and France during the 2009 H1N1 pandemic when lower H1N1 influenza cases coincided with rhinovirus outbreaks.^16,17^

Stringent NPIs may contribute to the significant reduction of all other non-influenza respiratory viral infections including RSV, hPMV, PIV1-3, adenovirus, enterovirus and rhinovirus. When the NPIs were relaxed after lockdown, the incidence of rhinovirus increased rapidly, a trend not seen with these other viruses. The mechanism behind this finding is unclear. High household transmission rates of rhinovirus, particularly between those contacts and their rhinovirus-infected children,^18^ plus a high proportion of asymptomatic infections^19^ may have contributed to this observation.

In the incoming autumn and winter of 2020 and 2021, many northern hemisphere temperate countries will have continuing COVID-19 circulation overlapping with the influenza season, resulting in increased burden on already stretched health systems. NZ’s experience strongly suggests that NPIs can greatly reduce the intensity of seasonal influenza and other respiratory viral infections. Continuation or strengthening of NPIs may, therefore, have positive impacts far beyond COVID-19 control. Even without these interventions, the severity of the 2020-2021 northern hemisphere influenza season remains uncertain. Both international and domestic air travel has been suggested as important drivers of influenza introduction and subsequent spread.^20^ It is possible that fewer seeding events from NZ and other southern hemisphere countries, from both reduced influenza activity and reduced air travel, may result in low influenza activity in these northern hemisphere countries during their incoming winter.

Our study has several limitations. Firstly, this is an observational study. Multiple simultaneous measures were applied depending on alert levels, making it difficult to understand the relative contribution of each of these measures. Secondly, during the COVID-19 laboratory response, some laboratories prioritised testing for COVID-19 and reduced testing for influenza and other respiratory viruses. Additionally, those samples ordered by clinicians for hospital inpatients and outpatients during normal clinical practices were based on clinician’s judgement, rather than a systematic sampling approach. This may result in selection bias. Thirdly, the government set up a number of community-based testing centres around the country to provide access to safe and free sampling for COVID-19. The usual flow and processes established for sentinel general practice based influenza-like illness surveillance may have been interrupted as many ILI patients would visit these centres instead of sentinel GP clinics. Additionally, national sentinel GP-based ILI surveillance requires swabbing from an ILI patient. This may contribute to the lower GP participation for this surveillance during the COVID-19 pandemic. These factors probably resulted in lower consultation and reporting and sample collection for sentinel ILI surveillance in 2020. However, the SARI surveillance system and SHIVERS-II&III cohorts operated as usual and showed the same apparent elimination of influenza virus circulation.

In conclusion, this observational study reported an unprecedented reduction in influenza and other important respiratory viral infections and the complete absence of an annual winter influenza epidemic, most likely due to the use of stringent nonpharmaceutical interventions (border restrictions, isolation and quarantine, social distancing and human behaviour changes). The data can inform future pandemic influenza preparedness and seasonal influenza planning for the northern hemisphere’s upcoming winter.

## Data Availability

All data is available upon request

## Methods

### Ethical approval

Ethical approval was obtained for the SHIVERS (including SARI and ILI surveillance), SHIVERS-II and III cohort studies from the NZ Northern A Health and Disability Ethics Committee (NTX/11/11/102). The ICD-coded influenza hospitalisation data and laboratory-based respiratory virus surveillance data are part of public health surveillance in NZ. It is conducted in accordance with the Public Health Act and thus ethics committee approval was not needed for collection or use of these data.

### Hospital-based severe acute respiratory illness (SARI) surveillance

The population-based hospital surveillance for severe acute respiratory illness (SARI) among residents (ca ∼1 million) of Central (Auckland District Health Board) and South (Counties Manukau District Health Board) Auckland was established in 2012.^21^ Research nurses reviewed daily records of all overnight acutely admitted inpatients to identify any inpatient with a suspected acute respiratory illness (ARI). They enrolled those patients with cough and history of fever (subjective fever or measured temperature ≥ 38°C) and onset within the past 10 days, defined by the World Health Organisation as SARI. A respiratory specimen (nasopharyngeal or nasal or throat swab) was collected and tested simultaneously for influenza and other respiratory viruses by real-time reverse transcription (RT) polymerase chain reaction (PCR) techniques:^22^ influenza virus, respiratory syncytial virus (RSV), rhinovirus, parainfluenza virus types 1-3, enterovirus, adenovirus, human metapneumovirus.

### Sentinel general practice-based influenza like-illness (ILI) surveillance

The population-based surveillance for influenza-like illness (ILI) among persons enrolled in sentinel general practices (∼90) who seek medical consultations has been in operation since 1990,^8^ usually covering ∼10% of the NZ population. The participating general practitioners and practice nurses assessed all consultation seeking patients. If a patient met the influenza-like illness (ILI) case definition: “an acute respiratory illness with a history of fever or measured fever of ≥ 38°C, and cough, and onset within the past 10 days, and requiring consultation in that general practice”, a respiratory specimen (nasopharyngeal or nasal or throat swab) was collected to test for influenza and other respiratory viruses.^21^ In 2020, sentinel general practice-based ILI surveillance was not operated in a usual way due to the COVID-19 response. Instead of visiting sentinel GPs for medical consultations, many ILI patients would visit one of the community-based testing centres. Additionally, national sentinel GP-based ILI surveillance requires swabbing from an ILI patient. This may contribute to less GP participation (18-57% of the usual participation rate over the winter period in 2020) in the COVID-19 pandemic situation. These factors would contribute to lower consultation, reporting and detection of influenza and other respiratory viruses compared with previous years.

### SHIVERS-II and WellKiwis cohort ILI surveillance

SHIVERS-II (the second iteration of the Southern Hemisphere Influenza and Vaccine Effectiveness Research and Surveillance programme) is a prospective adult cohort study in Wellington, NZ. The cohort study has been in operation since 2018 enrolling individuals aged 20-69 years, randomly selected from those healthy individuals listed in the general practice’s primarycare management system. In 2020, SHIVERS-II study staff followed these participants (∼1400) and monitored their ILIs and ARIs.

WellKiwis (i.e. SHIVERS-III) is a prospective Wellington infant cohort aiming to recruit 600 infant-mother pairs from Oct 2019-Sept 2022 (200 pairs a year) and follow them until 2026. In 2020, WellKiwis study staff followed up ∼80 infants and monitored their ILIs and ARIs.

During May-September 2020, SHIVERS-II and WellKiwis study staff sent weekly surveys to participants regarding their respiratory illness. Due to COVID-19, the ARI case definition in 2020 has changed to: “acute respiratory illness with fever or feverishness and/or one of following symptoms (cough, running nose, wheezing, sore throat, shortness of breath, loss of sense of smell/taste) with onset in the past 10 days.” The case definition for ILI during 2018-2020 was the same: acute respiratory illness with cough and fever/measured fever of ≥ 38□ and onset within the past 10 days. For those participants who met the case definition for ILI and ARI, research nurses visited the participant and take a nasopharyngeal or nasal or throat swab to test for influenza and other respiratory viruses and SARS-CoV-2.^23^

### International Classification of Diseases (ICD)-coded influenza hospitalisations

Hospitalisation data for ICD-coded influenza hospitalisations (ICD-10AM-VI codes (J09-J11) were extracted from the NZ Ministry of Health’s National Minimum Dataset by discharge date. In this dataset, patients who spent less than one day in a hospital are excluded. Influenza-related hospitalisations are conservatively taken to include only those cases where influenza was the principal diagnosis. Repeat hospital admissions were included because infection with a different influenza A sub-type or influenza B virus is possible.

### Laboratory-based surveillance

The laboratory-based surveillance for influenza and common respiratory viruses is carried out all-year-around by the NZ virus laboratory network consisting of the WHO National Influenza Centre (NIC) at ESR and six hospital laboratories in Auckland (2), Waikato, Wellington, Christchurch and Dunedin. This laboratory network tests specimens ordered by clinicians for hospital inpatients and outpatients during normal clinical practice (serving ∼70% of the NZ population). Sample collection is based on clinician’s judgement, rather than systematic sampling approach. This may result in selection bias. In addition, this laboratory network conducts testing for public health surveillance including SARI, ILI and SHIVERS-II and WellKiwis cohort surveillance.

### Data analyses

Study data were captured using REDCap electronic data capture tools.^24^ Analyses were performed in Stata 16.1 (StataCorp LLC).

The observed incidence rates of influenza-PCR-confirmed SARI or ILI or ARI were corrected each week to account for missed swabs from ILI cases by applying the influenza positivity rate of those tested to those non-tested (Corrected number of influenza-PCR-confirmed SARI or ILI or ARI events = Number of SARI or ILI or ARI x Actual number of influenza-PCR-confirmed SARI or ILI or ARI/Actual number of SARI or ILI or ARI swabs).

Based on SARI and ILI surveillance data from 2015-2019, the start of the annual influenza season and intensity level of the influenza epidemics was defined by using the Moving Epidemid Method (MEM).^25-27^ Briefly, MEM has three main steps: Step 1: for each season separately, the length of the epidemic period is estimated as the minimum number of consecutive weeks with the maximum accumulated percentage rates, splitting the season into three periods: a pre-epidemic, an epidemic, and a post-epidemic period; Step 2: MEM calculates the epidemic threshold as the upper limit of the 95% one-sided confidence interval of 30 highest pre-epidemic weekly rates, the n highest for each season taking the whole training period, where n = 30/number of seasons; Step 3: medium, high, and extra-ordinary intensity thresholds were estimated as the upper limits of the 40%, 90%, and 97.5% one-sided confidence intervals of the geometric mean of 30 highest epidemic weekly rates, the n highest for each season taking the whole training period, where n = 30/number of seasons. Five categories are used to set thresholds and define intensity level as no activity or below epidemic threshold, low (0-40%), moderate (40-90%), high (90-97.5%) and extra-ordinary (>97.5%) one sided confidence interval of the geometric mean.

The Cochran–Armitage test for trend analysis was performed for ICD-coded influenza hospitalisations and numbers of the reported virus detections.

Laboratory-based surveillance data and ICD-coded influenza hospitalisation data used the median weekly value to represent the reference period of 2015-2019. Median and interquartile ranges were calculated for the number of viruses reported during 2015-2019; Percentage of reduction = 1 - [No. virus (2020)/median no. virus (2015-2019)]%

## Acknowledgements

The SHIVERS-II project is funded by US National Institute of Allergy and Infectious Diseases (NIAID) (CEIRS Contract HHSN272201400006C). The WellKiwis (i.e. SHIVERS-III) project is funded by US-NIAID (U01 AI 144616). The SARI and ILI surveillance were funded by the NZ Ministry of Health during 2017-2020 and by US-Centers for Disease Control and Prevention (U01IP000480) during 2012-2016. The funding resource has no role in study design; collection, analysis or interpretation of data; writing of reports; nor decision to submit papers for publication.

SHIVERS-II & WellKiwis cohort study, SARI and ILI surveillance, led by the Institute of Environmental Science and Research (ESR), is a multi-centre and multi-disciplinary collaboration. The authors wish to thank SHIVERS collaborating organisations for their commitment and support: ESR, Auckland District Health Board (DHB), Counties Manukau DHB, Capital Coast DHB; Hutt Valley DHB; Regional Public Health; University of Auckland, University of Otago, WHO Collaborating Centre at St Jude Children’s Research Hospital in Memphis, USA. Wellington Maternity Health Professionals; Sentinel general practices; Local influenza coordinators within local Public Health Units; Participating hospital virology laboratories in ADHB, CMDHB, Waikato, Wellington, Dunedin and Christchurch;

## Authors’ contributions

All authors meet the International Committee of Medical Journal Editors criteria for authorship. QSH, RW, PT, BY, KD, AN, SJ, TD, NT, PCS, MB, HD, CCG, SJ, MGB, ST, AT, CW, SR, CM, TK designed and operationalised the SARI, ILI and/or SHIVERS-II&III cohort platforms; LJ, JR, WG, JB, JD, TM, ST, SR, FR, JB, CM, MD, AW, JG, AVDL, MB provided the testing and reporting; TJ, CE, WT, LE, MM, AM, SJ, LL, BW, HS, RG, KC, DC, SL, AS, TW, NA did the clinical data and samples collection and reporting and ensured operations; TW, QSH did the data analysis; QSH wrote the first draft of the manuscript. All authors contributed to the interpretation of the results, revision of the manuscript critically for intellectual content and have given final approval of the version to be published.

### The investigation team

#### The SARI research nurses at Auckland District Health Board (ADHB)

Alicia Stanley, Roxanne Buchanan, Pamela Muponisi, Marisa van Arragon, Medhawani Rao, Wendy Sullivan, Ellen Waymouth, Mapui Tangi, Emma Appleton, Leigh Smith, Debbie Aley, Bhamita Chand, Kathryn Haven, Stephnie Long, Julianne Brewer, Catherine McClymont, Claire Sherring, Miriam Rea, Shelley Barlow, Judith Tresidder, Judith Barry, Karen Bailey, Liz Walker, Medha Rao, Tracey Bushell

### The SARI research nurses at Counties Manukau District Health Board (CMDHB)

Shirley Laurence, Shona Chamberlin, Reniza Ongcoy, Kirstin Davey, Emilina Jasmat, Maree Dickson, Annette Western, Olive Lai, Sheila Fowlie, Faasoa Aupa’au, Louise Robertson; Emma Collis, Amanda Retter, Maricar Maminta, Paula Massey, Monica Jung, Renee Clark, Ashleigh Howan, Renee Clark, Bianca Underwood

### NZ virology laboratory network

Kamala Dullabh at ADHB; Lynell Hanley at Waikato DHB; Claire Tarring and Chor Ee Tan at Wellington SCL; Jennifer Fahey at Canterbury DHB;

### ESR National surveillance unit and laboratory

Namrata Prasad, Jill Sherwood, Erasmus Smit

## Disclaimer

The findings and conclusions in this report are those of the authors and do not necessarily represent the views of the US National Institute of Allergy and Infectious Diseases, US Department of Health and Human Services, the Institute of Environmental Science and Research (ESR) or any other collaborating organisations.

