## Supplementary Material for "Impact of the COVID-19 nonpharmaceutical interventions on influenza and other respiratory viral infections in New Zealand"

**Supplementary Figure 1.** Temporal distribution of influenza-like illness (ILI) from HealthStat general practice based surveillance in 2020 compared with the reference period of 2015-2019

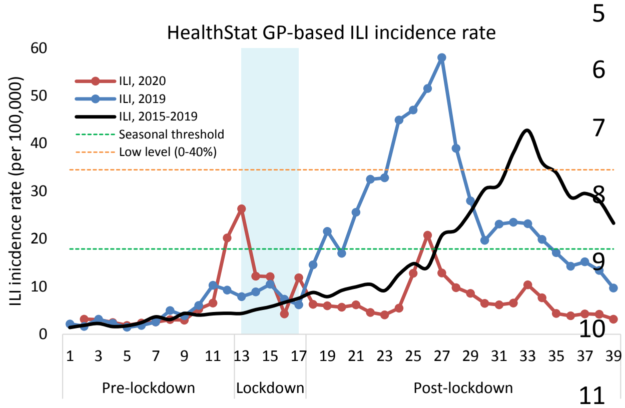


HealthStat general practice (GP) based ILI surveillance is based on a nationally representative random sample of approximately 100 general practices that code for ILI. The case definition used for ILI by HealthStat is "*acute upper respiratory tract infection, with abrupt onset of 2 or more symptoms from chills, fever, headache and myalgia*". This surveillance system monitors the number of people who consult GPs with an ILI. HealthStat is based on automated extracts from practice management computer systems. CBG Health Research Ltd provides this data to ESR on a weekly basis. HealthStat ILI surveillance does not include virological surveillance.

Supplementary Figure 1 showed that ILI incidence rates from HealthStat GP-based surveillance during May-September in 2020 were below the seasonal threshold (except week 26) compared to the reference period. This finding is consistent with national sentinel GP-based ILI surveillance and SHIVERS-II&III ILI/ARI surveillance as shown in Figure 1. Similar to the national sentinel GP-based ILI surveillance, some ILI patients from HealthStat may have been diverted to visit COVID-19 specific testing centres, resulting in lower than usual ILI reporting. However, HealthStat recorded higher ILI incidence rates than national sentinel GP-based ILI surveillance. As HealthStat does not have a virologic testing component, it may result in a similar level of GP participation compared to previous years. On the other hand, national sentinel GP-based ILI surveillance requires swabbing from an ILI patient. This may result in less GP participation (18-57% of the usual participation rate over the winter period in 2020) for this surveillance in the COVID-19 pandemic situation.

**Supplementary Figure 2**. Temporal distribution of non-influenza respiratory viral infections in 2020 compared with the reference period of 2015-2019

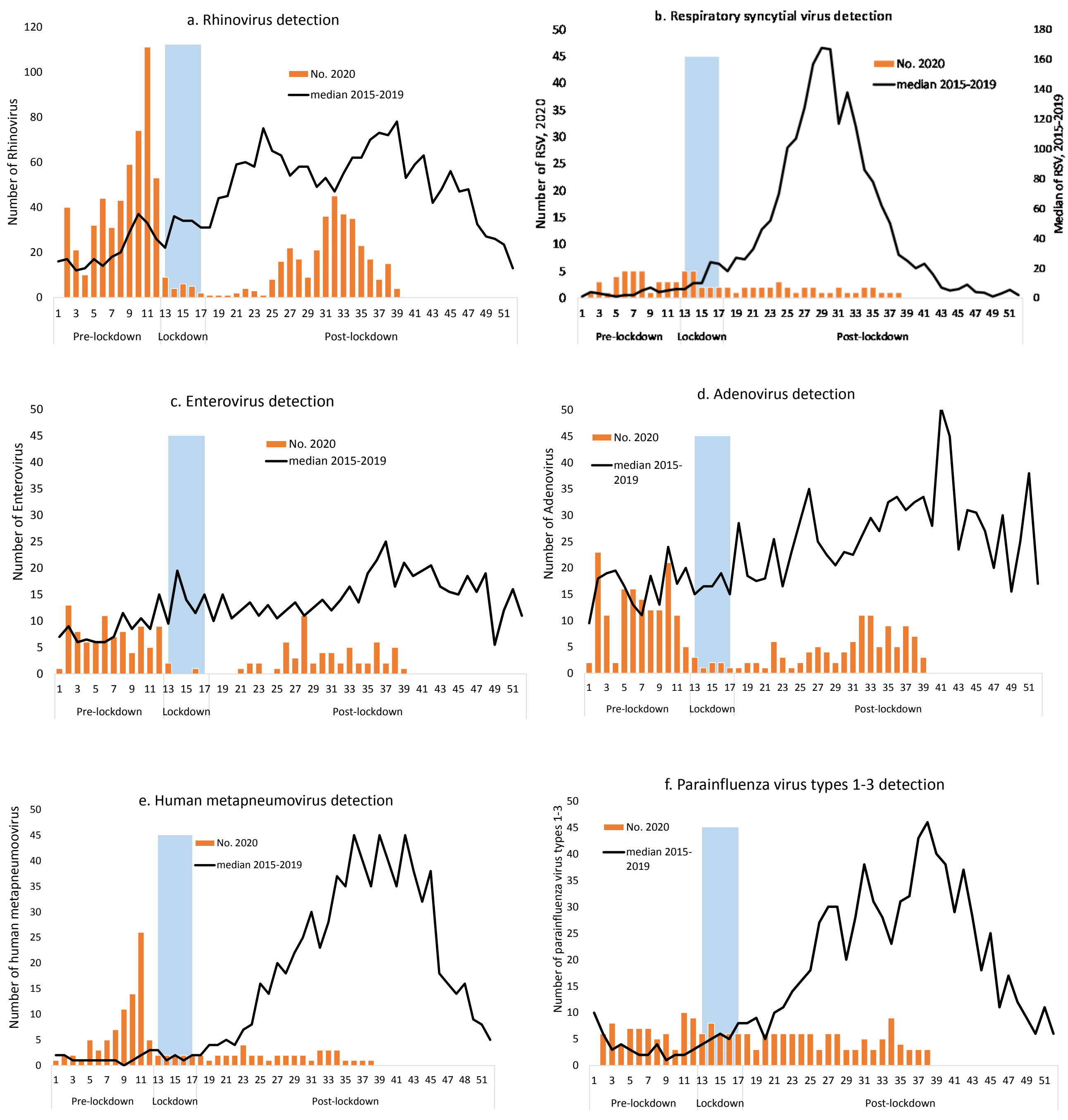

**Supplementary Figure 3**. Temporal distribution of rhinovirus infections among SHIVERS-II&III ARI cases and hospitalised SARI cases in 2020

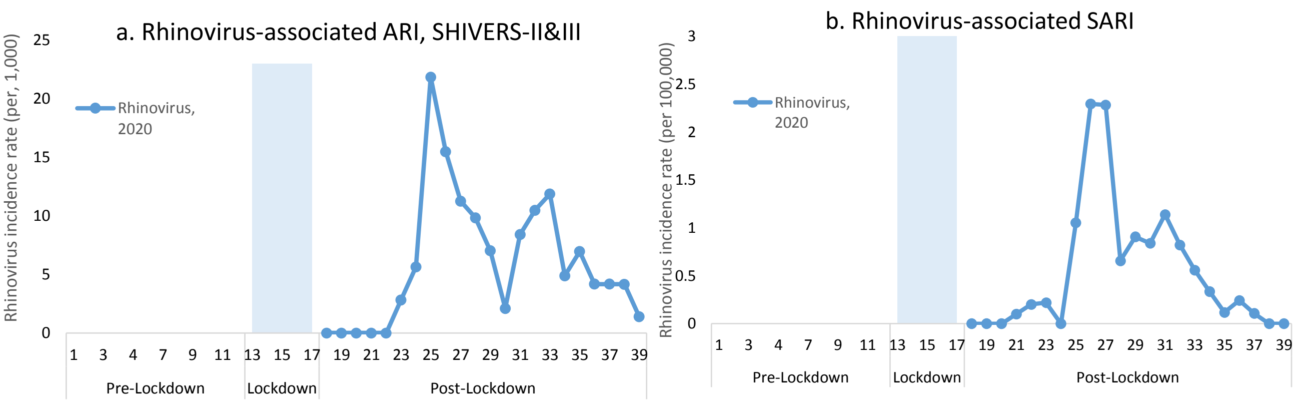
